# The African Breast Imaging Dataset for Equitable Cancer Care: Protocol for an Open Mammogram and Ultrasound Breast Cancer Detection Dataset

**DOI:** 10.1101/2025.08.26.25334483

**Authors:** Michael G. Kawooya, Richard Malumba, Alfred Jatho, Andrew Katumba, Joyce Nakatumba-Nabende, Denis Musinguzi, Simon Allan Achuka, Maruf Adewole, Udunna C Anazodo

**Affiliations:** Ernest Cook University; Uganda Cancer Institute; Makerere University; MAI Lab; McGill University

## Abstract

**Introduction:** Breast cancer is one of the most common cancers globally. Its incidence in Africa has increased sharply, surpassing that in high-income countries. Mortality remains high due to late-stage diagnosis, when treatment is less effetive. We propose the first open, longitudinal breast imaging dataset from Africa comprising point-of-care ultrasound scans, mammograms, biopsy pathology, and clinical profiles to support early detection using machine learning.

**Methods and Analysis:** We will engage women through community outreach and train them in self-examination. Those with suspected lesions, particularly with a family history of breast cancer, will be invited to participate. A total of 100 women will undergo baseline assessment at medical centers, including clinical exams, blood tests, and mammograms. Follow-up point-of-care ultrasound scans and clinical data will be collected at 3 and 6 months, with final assessments at 9 months including mammograms.

**Ethics and Dissemination:** The study has been approved by the Institutional Review Boards at ECUREI and the MAI Lab. Findings will be disseminated through peer-reviewed journals and scientific conferences.

## 1 Introduction

Breast cancer is among the most common types of cancer worldwide (1). Over the past 30 years, its incidence rate has increased globally due to compounding reproductive risk profiles such as advanced age at first birth, less breast feeding, oral contraceptives, behavioral risk factors, and improved cancer detection (2). While the incidence rate of breast cancer is reported to be higher in high income countries, rapid economic transition in Sub-Saharan Africa is driving an exponential rise in the number of new cases in the region (2; 3). It is projected that by 2040, the incidence rate of breast cancer in Sub-Saharan Africa will double (4).

Worse still, Black African women have the highest breast cancer mortality rate (5; 6). 50% of Black women diagnosed with breast cancer in Sub-Saharan Africa succumb to the disease within 5 years of diagnosis (2; 4). Meanwhile, in the United States of America that number is 20% and 10% for Black and White women respectively. Within Sub-Saharan Africa, 90% of white women in Namibia and 76% of mixed-race women in South Africa survive within 3 years of their diagnosis, compared with 56% Black Namibian women or 59% Black South African women(2). Uganda and Nigeria have one of the lowest survival rates, at 44-47% and 36%, respectively. The high mortality rates are largely attributed to late-stage diagnosis. This is caused by lack of access to prompt and early detection (2). 89% of the women in Uganda present with stage III or stage IV breast cancer (7). At this stage, the disease is more difficult to treat and the outcomes are poor.

The African Breast Cancer-Disparities in Outcomes Study (ABC-DO), a prospective study of 1,541 women newly diagnosed with breast cancer in Namibia, Nigeria, Uganda, and Zambia, found that socially disadvantaged women tended to be diagnosed at advanced stages of the disease (8). These included women with no formal education, poor breast cancer awareness, and younger age. Younger women who had been pregnant within the past three years were at particularly high risk, likely due to adverse hormone-related changes associated with pregnancy. More importantly, the study demonstrated that long distance to diagnostic facilities was associated with delay and late-stage diagnosis.

In a typical setting, women report to health centers after self-examination of their breast and armpit lymph nodes for lumps. They are then screened for suspected breast lesions using mammograms. Mammograms are low dose X-rays used as standard of care for breast care management (9). While frequent mammograms are encouraged for high risk patients, the cumulative ionizing radiation from X-ray exposure limits its use in frequent monitoring and further diagnostic investigations. When a lump or calcification is detected on a mammogram, an ultrasound scan is recommended for further investigation to determine if the tumor is benign or malignant. When a lump is detected on a mammogram, a complementary ultrasound is recommended to differentiate solid from cystic masses and to guide biopsy of the lesion. Ultrasound is a radiation free method. Malignancy is then confirmed by histology using biopsy. This clinical workflow is standard practice in high resource countries (10; 11). It has led to a significant increase in cancer detection especially at the earlier stages of the disease.

In Sub-Saharan Africa, mammograms are not readily available, so radiologists have largely turned to ultrasound as the primary screening method. However, when ultrasound scans are used independently, they have a lower positive predictive value (PPV) with biopsy confirmation(12). This is not surprising, as ultrasound has lower resolution than mammography and is more susceptible to intensity inhomogeneity, speckle noise, and a generally lower signal-to-noise ratio, making the detection of small tumors more challenging(13). As a result, accurate interpretation of breast ultrasound images heavily depends on the radiologist’s skill and experience. Unfortunately, there is a severe shortage of skilled radiologists in Sub Saharan Africa.

Machine learning–based tools for lesion segmentation and classification can support radiologists and help reduce diagnostic inequities(14; 15; 16; 17; 18). In resource-limited settings, ultrasound has proven effective at shifting breast cancer diagnoses from late to early stages, significantly improving survival rates(19; 20). By improving ultrasound’s positive predictive value (PPV), machine learning can make screening more accurate and reduce unnecessary biopsies.

However, in Sub-Saharan Africa, the lack of high-quality, expert-labeled breast ultrasound datasets poses a major barrier to applying machine learning in clinical settings. To date, there have been no known efforts to develop machine learning models for computer-aided screening, diagnosis, or risk assessment of breast cancer specifically in Sub-Saharan women. Compounding the issue, access to diagnostic imaging, including ultrasound, is severely limited. Breast imaging technologies are often restricted to a few centers in major cities, leaving rural and vulnerable populations without adequate diagnostic care.

To address these challenges, we propose creating a publicly available dataset of breast ultrasound images to support the development of machine learning models for early lesion detection. The dataset will be derived from a longitudinal study tracking suspicious cases over a nine-month period. We will collect conventional ultrasound scans from imaging centers, point-of-care ultrasound (POC-US) scans, mammograms, clinical data, and blood samples from women aged 35 to 49. The study will be conducted in Uganda and Nigeria, with each site contributing at least 50 cases. Qualified radiologists will annotate the imaging data with segmentation masks and diagnostic labels. The resulting dataset will enable training of segmentation and classification models aimed at improving early breast cancer detection in younger women across Sub-Saharan Africa.

## 2 Methods and Analysis

### 2.1 Study Design

Our central hypothesis is that with sufficient POC-US data, we can build models that match the positive predictive performance of mammograms in early detection of breast cancer. To test this hypothesis, we will conduct a longitudinal study of women between the age of 35-49 at Ernest Cook University in Kampala and Medical Artificial Intelligence Laboratory (MAI Lab) in Nigeria. We plan to develop machine learning models that will segment lesions and classify them according to the Breast Imaging Report and Imaging System (BIRADs)(21).

### 2.2 Patient population and sample size

The study will enroll female participants aged 35 to 49 years who report suspected breast lumps or related concerns and reside within 50 km of one of the following locations: Lagos, Nigeria; Kampala, Uganda; Nnewi, Nigeria; and Kano, Nigeria. The study period ran from January 2024 to December 2024. At each imaging center, participant samples will be stratified into three age groups: 15 women aged 35–39, 20 women aged 40–44, and 15 women aged 45–49.

Exclusion criteria include currently breastfeeding women, those undergoing treatment for any breast-related condition or any form of cancer, and those with previously diagnosed breast pathologies. Over the study duration, we anticipate collecting approximately 400 point-of-care ultrasound (POC-US) scans and 200 mammograms.

Ethical approval for the study has been obtained from the Institutional Review Boards at ECUREI and MAI Lab, and informed consent will be obtained from all participants.

### 2.3 Data Acquisition

Study participants will be recruited at public events such as community outreaches, concerts, religious gatherings, and other community settings. A letter of information containing full details about the project will be given to interested participants, to inform them of their roles. An informed consent form will be administered thereafter, followed by a guided interview to collect demographic information (age, height, weight), family history, pregnancy status, pregnancy history, and vitals about the participant. A POC-US will then be performed to identify normal and non-normal breast cases. The abnormal cases will be required to visit the principal study site (Crestview or Ernest Cook University and the Uganda Cancer Institute Radiology units) for further investigations. On arrival, the participants will be counseled on possible outcomes. A Conventional USS will be administered followed by a mammography to confirm whether the lesion is benign or malignant by a radiologist. Participants with malignant lesions will be referred to appropriate oncology centers for further diagnosis and treatment. Only participants with benign lesions will have blood work done. They will be followed up with vitals and POC-US at 2 different time points at 3-month intervals. The study will conclude with a mammography, vitals, conventional USS, and blood work at the 9-month mark. Figure 1 shows the data collection workflow.

**Figure 1.**
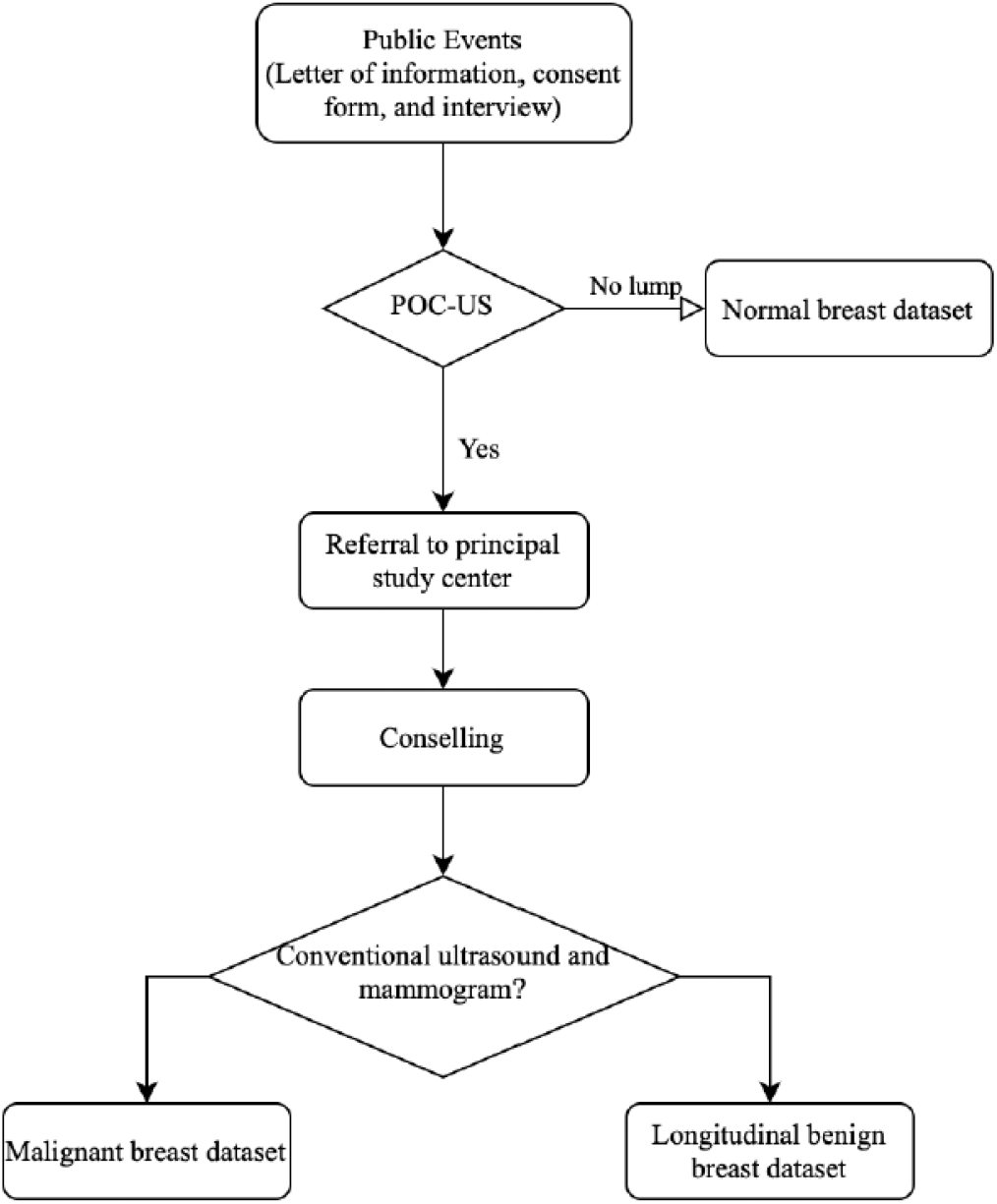
Mammogram and ultrasound data acquisition workflow.

The scanning procedure will vary based on the imaging modality and whether the patient presents with breast lesions. For breasts with no lesions, scans will include a 12 O’clock view and a complementary axillary view. For those with lesions, the same views will be obtained along with individual scans of each lesion at the point where its dimensions are largest. For mammography, four standard views will be captured: right and left medio-lateral oblique (MLO) and right and left cranio-caudal (CC) views.

### 2.4 Data Annotation

The dataset will provide both the overall assessment of the breast and information of the local level findings. The annotation of the dataset will follow the schema and lexicon of BI-RADS [21]. We will assess the quality of the images and categorize them into three classes: good, average and poor. At the breast level, the overall BI-RADS assessment categories and breast composition. There are seven BI-RADS assessment categories, namely BI-RADS 0 (need additional imaging or prior examinations), BIRADS 1 (negative), BI-RADS 2 (benign), BI-RADS 3 (probably benign), BI-RADS 4(suspicious), BI-RADS 5 (highly suggestive of malignancy), BI-RADS (known biopsy proven). Regarding the breast density for mammograms, its four categories are almost entirely fatty, scattered areas of fibroglandular, heterogeneous dense and extremely dense. For mammography findings, the list of findings includes mass, calcification, asymmetries, architectural distortion, and other associated features, namely suspicious lymph nodes, skin thickening, skin retraction, and nipple retraction. Each finding will be marked by a segmentation mask for localization. The masses will be classified by shape, margin, and density. The calcification will be classified by morphology and distribution.

For both POC-US and conventional ultrasound scans, the categories of breast tissue composition are homogeneous background echotexture-fat, homogeneous background echotexture-fibroglandular, and heterogeneous background echotexture. The lesions in the ultrasound scans will be classified into masses, calcifications, associated features, and special cases following BI-RADS. Masses will be categorized by shape, orientation, margin, echo pattern, and posterior features. Calcification will be categorized into calcification in a mass, calcification outside a mass and intraductal calcifications. Associated features will be categorized into architectural distortions, duct changes, skin changes, edema, vascularity, and elasticity assessments. The special cases include simple cyst, complicated cyst, clustered microcysts, mass in or on skin, foreign bodies including implants, lymph nodes-intramammary, lymph nodes-axillary, vascular abnormalities, post surgical fluid collection, fat necrosis and dilated lactiferous ducts..

The annotation of the images will be facilitated by a web-based annotation tool specifically designed for viewing and annotating medical images. The data will be labeled by a total of five radiologists, each labeling independently. All the radiologists hold healthcare professional certificates. They each have at least 5 years of experience reading soft tissue images. Each image will be labeled by two radiologists, one of which is a senior radiologist with experience of over 15 years in breast ultrasound. If there is no agreement between the two radiologists, a third tie radiologist, who is also a senior radiologist with over 15 years in breast ultrasound will act as a “tie-breaker.

After the annotation process, the breast-level categories and local annotations will be exported to JavaScript Object Notation (JSON) format. We will transform the data to comma-separated values (CSV) files so that it can easily be parsed. We will create a data dictionary to explain all the terms in the dictionary.

### 2.5 Data De-identification

The data will be strictly de-identified to protect patient privacy. All the meta-data in the DICOM images will be manually reviewed to ensure that all individually identifiable health information or PHI(22) of all the patients has been fully removed to meet data privacy regulations such as HIPAA(23). In addition, the image content of all mammograms will be manually reviewed case-by-case by human readers to ensure that no patient information remains.

## 3 Discussion

### 3.1 Implication and Future Directions

The dataset addresses a critical gap in global cancer diagnostics by providing the first African breast imaging data, collected using portable point-of-care devices in real-world low-resource settings. The inclusion of both mammography and ultrasound data, combined with longitudinal follow-up, will enable the development of AI tools capable of supporting screening, diagnosis, and disease monitoring even in regions without permanent imaging facilities or specialist radiologists.

Early detection improves treatment outcomes, as the disease is more manageable at its initial stages. In addition, these models could help address the shortage of radiologists by providing immediate, automated assessments at the point of care. By flagging abnormalities in real time, the models can reduce the workload on health workers and streamline the referral process. Importantly, they may also reduce reliance on mammography (which involves exposure to ionizing radiation) and thereby lower associated health risks. Furthermore, enhancing the positive predictive value of ultrasound scans could make early breast cancer detection more accessible, particularly in rural communities. Once the model has been developed, we plan to conduct a clinical trial to evaluate its accuracy and effectiveness in real-world settings.

However, the study does have limitations. The sample size will be relatively small compared to large, established datasets such as VinDR-Mammo(24), which may affect the generalizability of the models trained on this data. Additionally, the quality of images captured using POC-US devices may be variable and lower than conventional ultrasound systems, potentially impacting model performance.

### 3.2 Ethics and Dissemination

Upon completion of the study, we plan to publish our findings in peer-reviewed journals and present them at relevant medical and computer science conferences and workshops. To support further research, the dataset will be made available to other researchers under data use agreements. All shared data will be fully de-identified to protect patient privacy.

### 3.3 Strengths and Limitations

This study will produce the first ultrasound dataset specifically designed for cancer detection and classification in Black African women. Existing datasets largely consist of data from other regions, which may not reflect the unique breast tissue composition observed in this population. In contrast, this dataset will include comprehensive annotations that enable the development of a wide range of machine learning models to enhance breast cancer detection. For example, segmentation masks can be used to train models for both lesion segmentation and object detection, allowing for classification of lesions according to the seven BI-RADS categories. Additionally, the inclusion of both mammogram and ultrasound images will enable direct comparisons between the two modalities in Black African women, a group for whom mammography has shown limitations due to higher breast density. The longitudinal nature of the dataset will also support the development of models that predict disease progression or assess the impact of interventions over time.

## Data Availability

The data will be published after the study.

## References

[1] C. Fitzmaurice, C. Allen, R. Barber, L. Barregard, Z. Bhutta, H. Brenner, and G. B. of Disease Cancer Collaboration, “Global, regional, and national cancer incidence, mortality, years of life lost, years lived with disability, and disability-adjusted life-years for 32 cancer groups, 1990 to 2015: A systematic analysis for the global burden of disease study,” JAMA Oncology, 2017.

[2] P. Adeoye, “Epidemiology of breast cancer in sub-saharan africa,” in Breast Cancer Updates (S. Ş enözen and S. Emir, eds.), ch. 2, Rijeka: IntechOpen, 2023.

[3] D. Adeloye, O. Sowunmi, W. Jacobs, R. David, A. Adeosun, A. Amuta, S. Misra, M. Gadanya, A. Auta, M. Harhay, and K. Chan, “Estimating the incidence of breast cancer in africa: a systematic review and meta-analysis,” Journal of Global Health, vol. 8, no. 1, p. 010419, 2018.

[4] World Health Organization (WHO) and International Agency for Research on Cancer (IARC), “Breast cancer outcomes in sub-saharan africa.” https://www.iarc.who.int/wp-content/uploads/2021/03/IARC-Evidence-Summary-Brief-1.pdf,2021. IARC Evidence Summary Brief No.1.

[5] M. Galukande, H. Wabinga, and F. Mirembe, “Breast cancer survival experiences at a tertiary hospital in sub-saharan africa: A cohort study,” World Journal of Surgical Oncology, vol. 13, 2015.

[6] R. Sankaranarayanan, R. Swaminathan, H. Brenner, K. Chen, K. Chia, J. Chen, S. Law, Y.-O. Ahn, Y. Xiang, and B. Yeole, “Cancer survival in africa, asia, and central america: a population-based study,” The Lancet Oncology, vol. 11, no. 2, pp. 165–173, 2010.

[7] A. Gakwaya, J. Kigula-Mugambe, A. Kavuma, A. Luwaga, J. Fualal, J. Jombwe, M. Galukande, and D. Kanyike, “Cancer of the breast: 5-year survival in a tertiary hospital in uganda,” British Journal of Cancer, vol. 99, pp. 63–67, 2008.

[8] K. Togawa, B. Anderson, M. Foerster, M. Galukande, A. Zietsman, J. Pontac, A. Anele, C. Adisa, G. Parham, L. Pinder, F. McKenzie, J. Schüz, I. Santos-Silva, and V. McCormack, “Geospatial barriers to healthcare access for breast cancer diagnosis in sub-saharan african settings: The african breast cancer–disparities in outcomes cohort study,” International Journal of Cancer, vol. 148, pp. 2212–2226, 2021.

[9] M. Zeeshan, B. Salam, Q. Khalid, S. Alam, and R. Sayani, “Diagnostic accuracy of digital mammography in the detection of breast cancer,” Cureus, 2018.

[10] K. Kerlikowske, L. Ichikawa, D. Miglioretti, D. Buist, P. Vacek, R. Smith-Bindman, B. Yankaskas, P. Carney, and R. Ballard-Barbash, “Longitudinal measurement of clinical mammographic breast density to improve estimation of breast cancer risk,” Journal of the National Cancer Institute, vol. 99, no. 5, pp. 386–395, 2007.

[11] E. Devolli-Disha, S. Manxhuka-Kërliu, H. Ymeri, and A. Kutllovci, “Comparative accuracy of mammography and ultrasound in women with breast symptoms according to age and breast density,” Bosnian Journal of Basic Medical Sciences, vol. 9, p. 131, 2009.

[12] W. Berg, A. Bandos, E. Mendelson, D. Lehrer, R. Jong, and E. Pisano, “Ultrasound as the primary screening test for breast cancer: Analysis from acrin 6666,” Journal of the National Cancer Institute, vol. 108, p. 367, 2016.

[13] O. Sogunro, C. Cashen, S. Fakir, J. Stausmire, and N. Buderer, “Detecting accurate tumor size across imaging modalities in breast cancer,” Breast Disease, vol. 40, no. 3, pp. 177–182, 2021.

[14] Q. Zhang, Y. Xiao, W. Dai, J. Suo, C. Wang, J. Shi, and H. Zheng, “Deep learning based classification of breast tumors with shear-wave elastography,” Ultrasonics, vol. 72, pp. 150–157, 2016.

[15] A. Becker, M. Mueller, E. Stoffel, M. Marcon, S. Ghafoor, and A. Boss, “Classification of breast cancer in ultrasound imaging using a generic deep learning analysis software: A pilot study,” British Journal of Radiology, vol. 91, no. 1089, p. 20170576, 2018.

[16] S. Han, H.-K. Kang, J.-Y. Jeong, M.-H. Park, W. Kim, W.-C. Bang, and Y.-K. Seong, “A deep learning framework for supporting the classification of breast lesions in ultrasound images,” Physics in Medicine & Biology, vol. 62, pp. 7714–7728, 2017.

[17] R. Almajalid, J. Shan, Y. Du, and M. Zhang, “Development of a deep-learning based method for breast ultrasound image segmentation,” in 2018 17th IEEE International Conference on Machine Learning and Applications (ICMLA), (Orlando, FL, USA), pp. 1103–1108, IEEE, 2018.

[18] U. Anazodo, M. Adewole, and F. Dako, “Ai for population and global health in radiology,” Radiology: Artificial Intelligence, vol. 4, no. 4, p. 220107, 2022.

[19] F. Nandan and B. Alladin, “The role of ultrasound as a diagnostic tool for breast cancer in the screening of younger women,” Journal of Medical Diagnostic Methods, vol. 7, 2018.

[20] R. Sood, A. Rositch, D. Shakoor, E. Ambinder, K. Pool, E. Pollack, D. Mollura, L. Mullen, and S. Harvey, “Ultrasound for breast cancer detection globally: A systematic review and meta-analysis,” JCO Global Oncology, vol. 5, pp. 1–17, 2019.

[21] C. D’Orsi, E. Sickles, E. Mendelson, and E. Morris, ACR BI-RADS Atlas: Breast Imaging Reporting and Data System. Reston, Virginia: American College of Radiology, 2013.

[22] S. Isola and Y. Al Khalili, Protected Health Information. Treasure Island (FL): StatPearls Publishing, 2025. [Updated 2023 Jan 30].

[23] US Department of Health and Human Services, “Summary of the hipaa privacy rule.” https://www.hhs.gov/hipaa/for-professionals/privacy/laws-regulations/index.html, 2003. Accessed 2025-06-16.

[24] H. Nguyen, H. Nguyen, H. Pham, K. Lam, L. Le, M. Dao, and V. Vu, “Vindr-mammo: A large-scale benchmark dataset for computer-aided diagnosis in full-field digital mammography.” https://arxiv.org/abs/2203.11205, 2023.

